# From ‘Negative’ Trial to Positive Clinical Impact: Emulating WARCEF While Accounting for Selection Bias in Trial Timing

**DOI:** 10.1101/2025.03.19.25324271

**Authors:** Xiaodi Li, Sivaraman Rajaganapathy, Xinyue Hu, Jingna Feng, Jianfu Li, Yue Yu, Phil Fiero, Soulmaz Boroumand, Richard Larsen, Xiaoke Liu, Cui Tao, Nansu Zong

## Abstract

While randomized controlled trials (RCTs) guide clinical practice, their completion may also influence real-world treatment patterns. We investigated whether the outcomes of trial emulation differ before and after the publication of the WARCEF (Warfarin versus Aspirin in Reduced Cardiac Ejection Fraction) randomized controlled trial. We emulated the WARCEF trial using EHR data from the Mayo Clinic Platform, comparing Warfarin and Aspirin in patients with heart failure and reduced ejection fraction (HFrEF). Analyses were stratified by the WARCEF completion date (July 2014), using both intention-to-treat (ITT) and per-protocol (PP) frameworks. For the ITT analysis, the cohort size before 2014 consisted of 4,579 patients on Aspirin and 93 on Warfarin, while after 2014, the cohort included 13,599 patients on Aspirin and 493 on Warfarin. For the PP analysis, the cohort size before 2014 consisted of 4,314 patients on Aspirin and 86 on Warfarin, while after 2014, the cohort included 3,373 patients on Aspirin and 99 on Warfarin. No significant treatment difference was observed before July 2014, 1.3961 (95% CI: 0.696 – 2.802, p = 0.3477) and 1.1572 (95% CI: 0.559 – 2.397, p = 0.6945) for ITT and PP analytic approaches respectively, consistent with the findings of the WARCEF trial. However, after trial completion, Warfarin was associated with increased risk, 2.039 (95% CI: 1.412 – 2.945, p < 0.001) and 3.940 (95% CI: 1.376 – 11.286, p = 0.0107) for ITT and PP analysis respectively. These changes suggest that the WARCEF results and subsequent guideline updates influenced prescribing patterns and patient selection, thereby altering the observed treatment effects. Trial completion and guideline shifts can materially affect real-world emulation outcomes. Ignoring temporal changes in clinical practice may obscure causal inference and overstate or understate treatment effects in emulation studies. The publication of the WARCEF trial results may have influenced clinical decision-making, leading physicians to prescribe Warfarin or Aspirin differently based on trial findings. This shift in treatment patterns can introduce selection bias in real-world data, as patient characteristics associated with each treatment may no longer be randomly distributed after the trial. For accurate trial emulation, it is essential to ensure that the patient cohort aligns precisely with the trial environment, particularly with respect to the trial’s completion date.

## 1. Introduction

Randomized controlled trials (RCTs) are widely regarded as the gold standard for establishing causal relationships in clinical research due to their ability to minimize bias and confounding [1]. Positive RCT findings often lead to updates in clinical guidelines and changes in medical practice [2, 3]. However, the impact of negative or inconclusive RCTs on clinical practice is less clear. Studies have shown that while successful trials can rapidly shift treatment standards, negative results do not consistently lead to practice changes, and ineffective interventions may persist despite emerging evidence [4, 5]. Our study uses real-world data while strictly adhering to the original trial criteria and time frame and reveals clear differences between data collected before and after the trial’s completion date.

The Warfarin versus Aspirin in Reduced Cardiac Ejection Fraction (WARCEF) trial was designed to determine whether Warfarin or Aspirin is more effective in preventing death and ischemic stroke among patients with heart failure and reduced ejection fraction (EF ≤ 35%). Although Warfarin has shown benefits in ischemic heart disease and atrial fibrillation, its role in heart failure patients without atrial fibrillation remained unclear. In this randomized, double-blind, multicenter trial enrolling 2,305 patients, WARCEF found no significant difference between Warfarin and Aspirin for the composite outcome of death (hazard ratio [HR] 1.01; 95% CI, 0.85 to 1.20; p = 0.91), with Warfarin reducing ischemic stroke risk but increasing major bleeding events. These findings are consistent with results from retrospective analyses [6-8] and other randomized trials [9-11], which similarly showed no significant reduction in thromboembolic events with Warfarin in heart failure patients.

Clinical guidelines evolve with emerging trial evidence, influencing both practice and future research. Prior to the WARCEF trial, recommendations on anticoagulation in heart failure with reduced ejection fraction (HFrEF) and sinus rhythm were largely extrapolated from atrial fibrillation populations, as reflected in the 2010 ESC Guidelines [12]. WARCEF addressed this gap by comparing Warfarin and Aspirin in HFrEF patients without atrial fibrillation, ultimately reporting no overall benefit due to offsetting reductions in ischemic stroke and increases in major bleeding. Following its publication, updated guidelines, including the 2020 [13] and 2024 ESC [14] and 2022 AHA/ACC/HFSA [15], recommended against routine anticoagulation in this population unless clear indications exist. Together with findings from the COMMANDER HF trial, WARCEF reshaped guideline recommendations and indirectly influenced cohort definitions, treatment comparisons, and outcome expectations in subsequent trial emulations.

This study seeks to emulate the WARCEF trial using real-world observational data from the Mayo Clinic Platform (MCP). Our goal is to evaluate the comparative effectiveness of anticoagulation strategies, specifically Warfarin versus Aspirin, in patients with HFrEF. To strictly replicate the original trial design, we rigorously aligned our inclusion and exclusion criteria with those of the WARCEF trial and confined our pre-trial analysis to data collected prior to the trial’s formal completion in 2014. By comparing patient outcomes before and after this landmark date, we aim to determine whether the publication of the WARCEF trial influenced real-world treatment patterns and corresponding outcomes. This approach not only provides insight into the external validity of the trial findings but also helps assess the translational impact of RCTs on clinical practice over time.

Our method makes a threefold contribution: (1) demonstrating the impact of trial completion on the effectiveness of medication in improving patient outcomes; (2) demonstrating the feasibility of RCT emulation using real-world data (RWD) within MCP, ensuring that replication methods can be systematically applied in clinical settings; (3) addressing methodological challenges in observational research, such as data processing complexities, variable selection, confounder balancing, and adapting trial protocols for structured and reproducible replication.

## 2. Results

### 2.1 Study Details

Table 1 summarizes the WARCEF trial, and the corresponding patient cohort identified from the MCP, including patient counts across different stratification methods and time periods. We conducted analyses using both ITT (Intention-To-Treat) and PP (Per-Protocol) approaches and further divided the cohort into two groups: before and after July 2014.

**Table 1.**
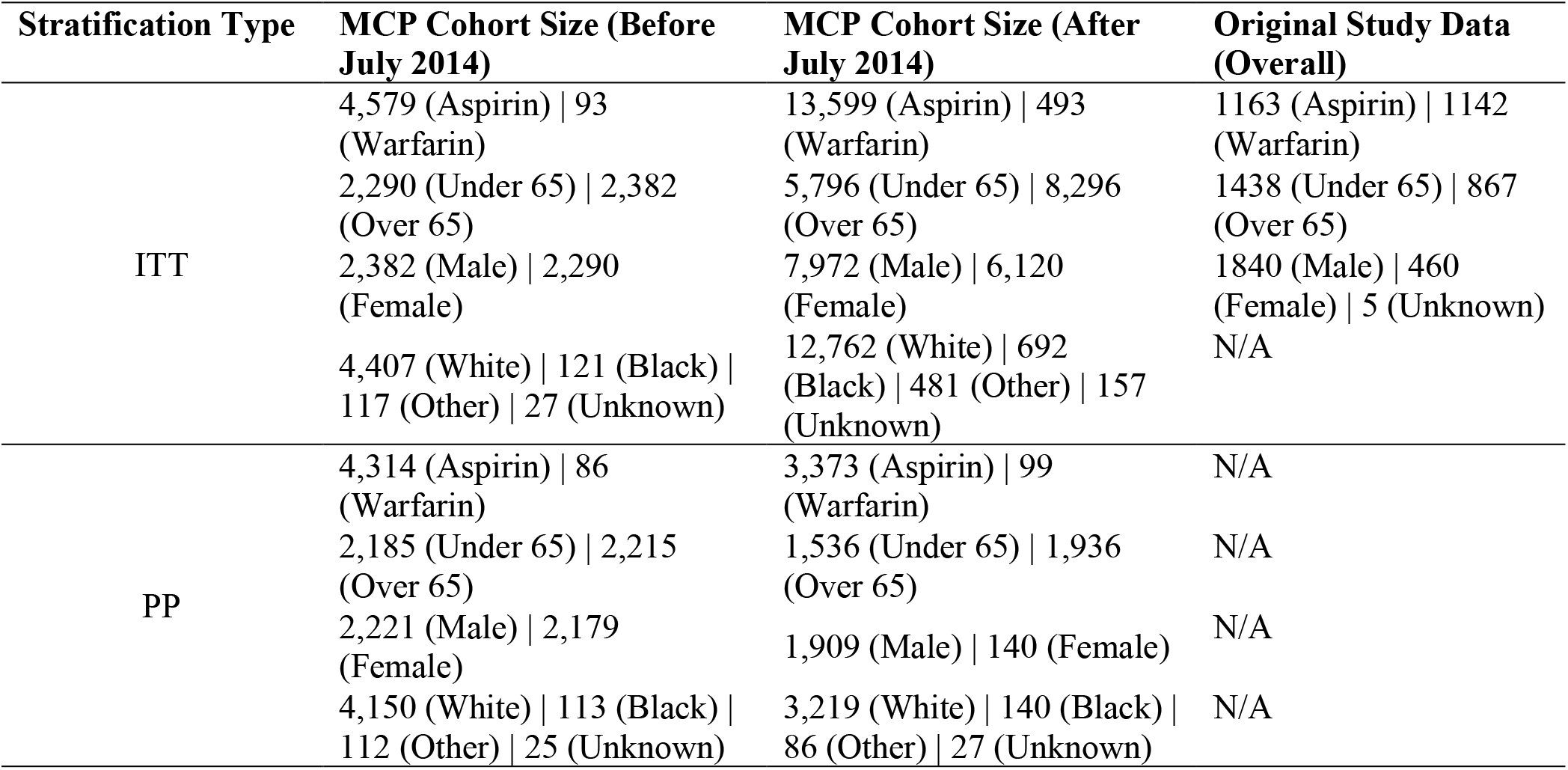
Study Summaries.

Throughout the process, we devoted substantial effort to variable selection. In collaboration with physicians, we carefully reviewed and discussed which variables to include or exclude, guided by the official inclusion and exclusion criteria outlined on the trial’s website. We strictly included variables deemed essential and excluded those considered clear disqualifiers, such as chronic or paroxysmal atrial fibrillation (AF), presence of a mechanical valve, and age under 18. Remaining variables were treated as potential confounders. After applying these criteria, we filtered the initial cohort from 38,172 patients down to 18,765. To closely emulate the WARCEF trial environment, we divided our dataset into two periods: before July 2014 and after July 2014. The pre-July 2014 cohort corresponds to the original trial period, capturing patients before the trial results were disseminated. The post-July 2014 cohort includes patients after the trial’s completion, reflecting the potential influence of WARCEF findings on real-world clinical practice.

### 2.2. Detailed Results

We assessed the mortality outcomes by comparing real-world emulated cohorts before and after the completion of the WARCEF trial in July 2014. Hazard ratio analyses, including key clinical risk factors, were conducted across the full study population without separately distinguishing between ITT and PP frameworks. As shown in Table 2, before July 2014, the comparison between Warfarin and Aspirin showed no statistically significant difference in mortality. The hazard ratios were 1.3961 (95% CI: 0.696 – 2.802, p = 0.3477) and 1.1572 (95% CI: 0.559 – 2.397, p = 0.6945) for ITT and PP analytic approaches respectively, suggesting a trend toward higher mortality with Warfarin but without reaching conventional significance. For the ITT analysis before July 2014, significant risk factors included Black race (HR = 0.0476, p = 0.0273), presence of contraindications (HR = 0.1312, p = 0.0087), pregnancy (HR = 0.0247, p = 0.0014), and severe liver impairment (HR = 3.5311, p = 0.0313). In the PP analysis during the same period, the presence of contraindications remained a significant risk factor (HR = 0.0715, p < 0.001).

**Table 2.** Hazard Ratio for Mortality.

| ITT Before July 2014 |  |  | ITT After July 2014 |  |  |
| --- | --- | --- | --- | --- | --- |
| Medication (Warfarin) |  |  | Medication (Warfarin) |  |  |
| Hazard Ratio |  | p-value | Hazard Ratio |  | p-value |
| 1.3961 (0.696 – 2.802) |  | 0.3477 | 2.039 (1.412 – 2.945) |  | <0.001 |
| Risk Factors |  |  | Risk Factors |  |  |
| RF | Hazard Ratio | p-value | RF | Hazard Ratio | p-value |
| Race (Black) | 0.0476 (0.003 – 0.711) | 0.0273 | Age (Over 65) | 1.662 (1.112 – 2.486) | 0.0133 |
| Contraindications (Present) | 0.1312 (0.029 – 0.599) | 0.0087 | Gender (Female) | 1.472 (1.007 – 2.153) | 0.0461 |
| Pregnancy (Present) | 0.0247 (0.003 – 0.241) | 0.0014 | Alcohol (Former) | 2.139 (1.324 – 3.454) | 0.0019 |
| Severe liver impairment (Present) | 3.5311 (1.120 – 11.135) | 0.0313 | Modified Rankin score (Below 4) | 1.858 (1.273 – 2.711) | 0.0013 |
|  |  |  | ACE inhibitor use (Present) | 0.264 (0.127 – 0.549) | <0.001 |
| PP Before July 2014 |  |  | PP After July 2014 |  |  |
| Medication (Warfarin) |  |  | Medication (Warfarin) |  |  |
| Hazard Ratio |  | p-value | Hazard Ratio |  | p-value |
| 1.1572 (0.559 – 2.397) |  | 0.6945 | 3.940 (1.376 – 11.286) |  | 0.0107 |
| Risk Factors |  |  | Risk Factors |  |  |
| RF | Hazard Ratio | p-value | RF | Hazard Ratio | p-value |
| Contraindications (Present) | 0.0715 (0.015 – 0.333) | <0.001 | Age (Over 65) | 5.495 (2.121 – 14.240) | <0.001 |
|  |  |  | Race (Black) | 34.053 (4.681 – 247.721) | <0.001 |
|  |  |  | Smoking (Heavy) | 0.198 (0.054 – 0.725) | 0.0145 |
|  |  |  | Alcohol (Yes) | 3.117 (1.131 – 8.591) | 0.0279 |
|  |  |  | NSAID (Present) | 5.494 (1.796 – 16.801) | 0.0028 |

In contrast, as shown in Table 2, after July 2014, Warfarin was consistently associated with significantly higher mortality compared to Aspirin. The hazard ratios increased to 2.039 (95% CI: 1.412 – 2.945, p < 0.001) and 3.940 (95% CI: 1.376 – 11.286, p = 0.0107) for ITT and PP analysis respectively, indicating a statistically significant increase in mortality associated with Warfarin. For the ITT analysis after July 2014, significant risk factors included age over 65 (HR = 1.662, p = 0.0133), female gender (HR = 1.472, p = 0.0461), former alcohol use (HR = 2.139, p = 0.0019), modified Rankin score below 4 (HR = 1.858, p = 0.0013), and ACE inhibitor use, which was associated with decreased risk (HR = 0.264, p < 0.001). In the PP analysis after July 2014, Warfarin remained significantly associated with elevated risk (HR = 3.940, 95% CI: 1.376–11.286, p = 0.0107). Significant risk factors in this group included age over 65 (HR = 5.495, p < 0.001), Black race (HR = 34.053, p < 0.001), heavy smoking (HR = 0.198, p = 0.0145), alcohol use (HR = 3.117, p = 0.0279), and NSAID use (HR = 5.494, p = 0.0028).

For the combined cohort including both pre- and post-July 2014 data, the ITT hazard ratio was 2.136 (95% CI: 1.471 – 3.101, p < 0.001), and the PP hazard ratio was 3.928 (95% CI: 1.8775 – 8.22, p < 0.001). These values closely mirror those observed in the post-July 2014 subgroup, suggesting that the overall cohort was heavily influenced by data collected after the trial’s completion. This highlights the potential for post-trial changes in treatment practices to dominate aggregated analyses. It further underscores the need to consider temporal stratification when interpreting real-world evidence.

These findings suggest that while no significant treatment difference was observed prior to WARCEF trial completion, a clear and statistically significant survival benefit favouring Aspirin emerged afterward. This temporal shift underscores the influence of clinical trial completion and dissemination on real-world treatment effectiveness. It also highlights how treatment effects may evolve over time in response to changing clinical practices, awareness, and decision-making processes. The divergence between pre- and post-trial periods suggests that real-world evidence is not static and underscores the importance of time-sensitive analyses in trial emulation studies. These results reinforce the need to consider temporal context when evaluating treatment outcomes and interpreting the broader impact of clinical trials beyond their publication.

As shown in Figure 1, both the ITT and PP analyses indicate that Warfarin is associated with higher mortality compared to Aspirin. While the difference between the two treatments was not statistically significant before 2014, a significant divergence emerges after 2014, with Aspirin showing a clear survival advantage.

**Figure 1.**
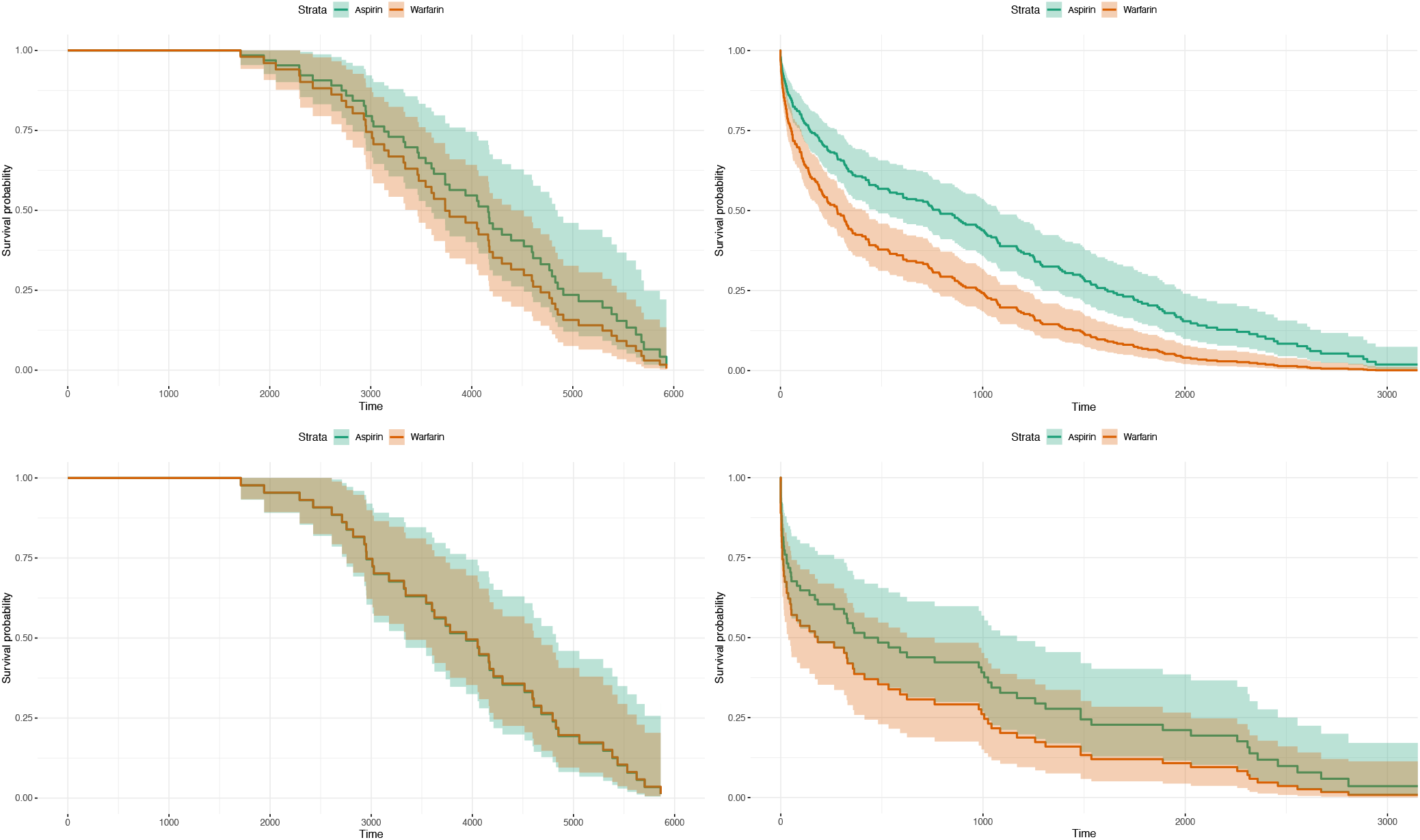
Survival Curves Stratified by Analysis Type and Time Period. This figure presents Kaplan–Meier survival curves comparing Aspirin (green) and Warfarin (orange) under both ITT and PP analyses. The plots are organized as follows: the top row corresponds to ITT, and the bottom row corresponds to PP. The left column shows data from before 2014, and the right column shows data from after 2014. Prior to 2014, no significant difference in survival was observed between the two treatments in either ITT or PP analyses. However, after 2014, a clear separation emerges between the curves, indicating a statistically significant survival benefit for patients treated with Aspirin. Across all stratifications, Aspirin consistently demonstrates lower mortality risk compared to Warfarin, particularly in the post-2014 period.

Figure 2 presents the estimated coefficients for clinical variables identified as confounders in the ITT analysis conducted after the WARCEF trial’s completion. The bar plot reveals that lifestyle-related factors such as light smoking and former alcohol use are among the strongest predictors of adverse outcomes, suggesting that behavioural history may play a more prominent role in shaping risk profiles in the post-trial population. Additionally, variables related to treatment, such as use of antiplatelet agents and Warfarin, show meaningful positive associations, potentially reflecting both medication effects and changes in prescribing patterns after 2014. On the other hand, variables with negative coefficients, such as intravenous heparin use, may signal protective influences or indicate selection mechanisms that emerged in real-world practice. Together, these findings demonstrate the heterogeneous impact of clinical characteristics on patient outcomes following trial dissemination and underscore the importance of carefully accounting for confounding when emulating trial results in post-trial data.

**Figure 2.**
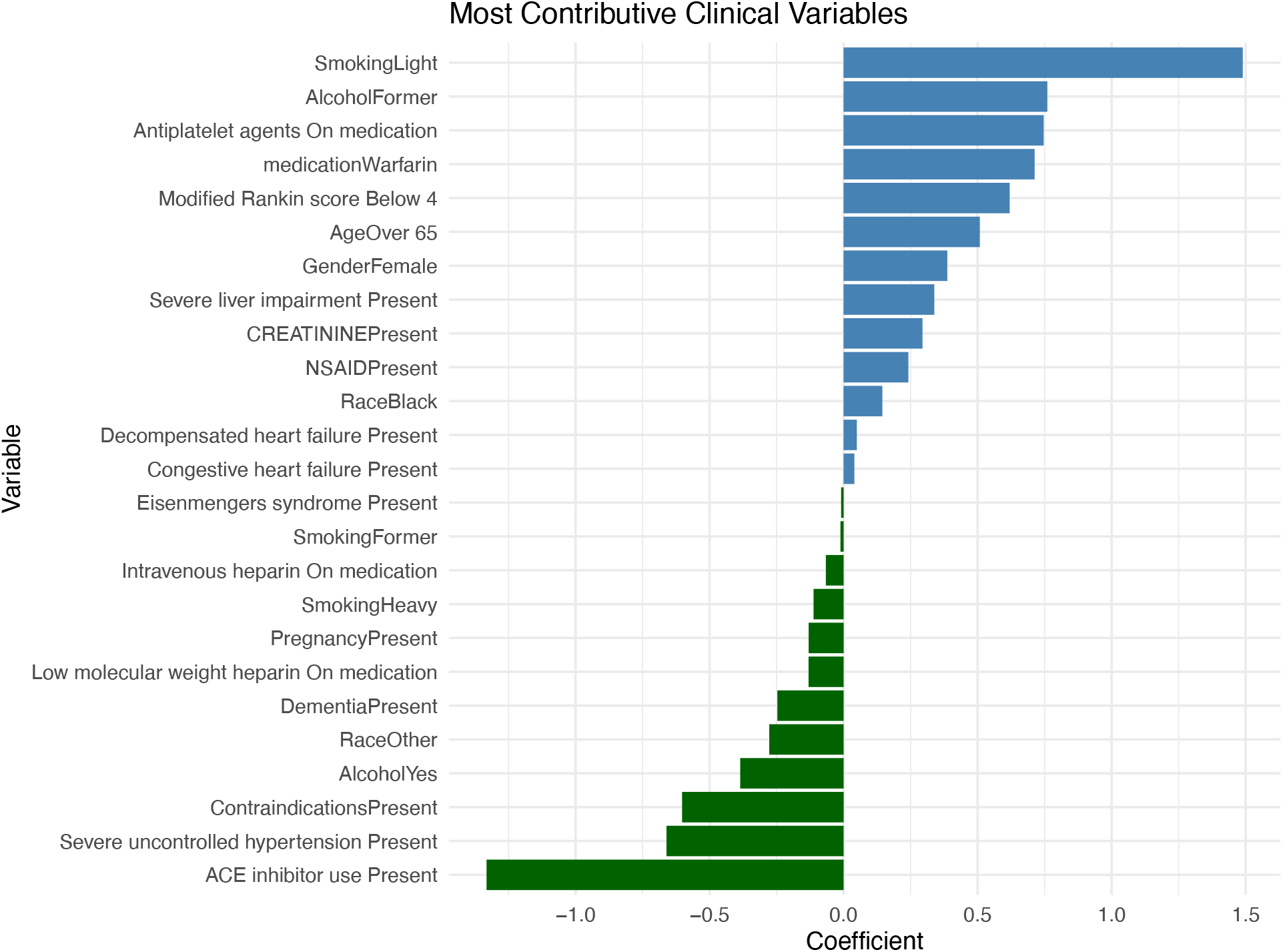
Confounders Coefficient of ITT after 2014. This figure illustrates the most contributive clinical variables influencing outcomes in the ITT analysis conducted after 2014. The horizontal bar plot presents variable names along the y-axis and their corresponding coefficient values along the x-axis. Positive coefficients reflect stronger associations with increased outcome probability or risk, while negative coefficients indicate potential protective effects. The most influential positive contributors include “SmokingLight”, and “AlcoholFormer”, which appear at the top of the chart. These variables are associated with elevated outcome risk and likely play a key role in driving adverse patient trajectories. Other important positive features include “Antiplatelet agents On medication” and “medicationWarfarin”, further emphasizing the significance of treatment-related and cardiovascular factors. Toward the middle of the distribution, variables like “CREATININEPresent” and “NSAIDPresent” show smaller positive coefficients, suggesting a moderate influence on outcomes. Meanwhile, several variables exhibit negative coefficients, indicating potentially protective associations. These include “PregnancyPresent” and “Intravenous heparin On medication”. Notably, clinical conditions such as “SmokingFormer” and “Eisenmengers syndromePresent” appear at the lower end with the most negative coefficients. These variables show an inverse relationship with the outcome in this cohort, possibly reflecting treatment effects, selection bias, or post-2014 shifts in clinical practice. Overall, this visualization highlights the varying degrees to which individual clinical variables impact patient outcomes post- 2014, with smoking behaviour, antihypertensive use, and anticoagulant medication emerging as particularly significant factors.

Supplement Figure 1, 2, and 3 in the Appendix provide additional coefficient plots for the ITT analysis before 2014, as well as the PP analyses conducted before and after 2014.

## 3. Discussion

This study demonstrates the feasibility of using RWD from the Mayo Clinic Platform to emulate RCTs and assess treatment effects in patients with HFrEF. By replicating the WARCEF trial design, we found that treatment effects differed substantially before and after the trial’s completion in 2014: no significant difference between Warfarin and Aspirin was observed prior to 2014, whereas a statistically significant difference emerged in the post-trial period. This suggests that dissemination of the WARCEF findings likely influenced real-world prescribing patterns and clinical decision-making. The results highlight the potential of trial emulation to capture temporal shifts in practice, revealing how evidence from RCTs may be translated into patient care. Our findings underscore the importance of aligning real-world analyses with trial timelines and clinical context to better understand treatment dynamics and adoption in practice.

Although the WARCEF trial reported a neutral primary outcome [11], its publication likely amplified its visibility and influence on clinical decision-making. High-profile trials, even those with negative or neutral results, can shift practice when addressing key therapeutic uncertainties. Supporting this, Homma et al. [16] showed that among patients under 60 years old, Warfarin was associated with improved outcomes compared to Aspirin, while older patients saw no such benefit, indicating the interpretation clinicians may apply following publication. Furthermore, quality of anticoagulation control, measured by time in therapeutic range (TTR), was found to be a critical modifier of treatment efficacy: higher TTRs were significantly associated with better outcomes in Warfarin-treated patients [17]. These findings underscore how post-trial analyses can reinforce or refine clinical interpretations, further affecting treatment patterns.

Nonetheless, broader shifts in heart failure management, such as evolving clinical guidelines, new therapeutic options, and changes in care delivery, may also contribute to the observed treatment effect differences over time. These systemic developments can confound real-world analyses and must be considered alongside the influence of individual trial findings. Real-world emulations must therefore be contextualized within the broader landscape of clinical practice, which is continually evolving. Our findings are preliminary but underscore the importance of further research to understand how and when trial publications impact clinical behavior, particularly across diverse health systems and patient populations. In addition, accurate trial emulation demands rigorous adherence to the original trial’s eligibility criteria, intervention definitions, and study timeline. Ensuring that the emulated patient cohort aligns precisely with the trial period is critical for producing valid and interpretable results.

Through the trial emulation process, we use Cox model [18] to do the survival analysis. We found that the Cox model’s outputs are highly sensitive to how covariates are specified, particularly the selection and treatment of confounding variables. For instance, defining ACE inhibitor use as a strict inclusion criterion resulted in a HR below 1, suggesting a protective effect, whereas treating it as a confounder produced an HR above 1, implying potential harm. This discrepancy underscores a crucial methodological challenge: the outcome of emulated trials can vary substantially based on how variables are handled. To mitigate such inconsistencies, there is a pressing need for a principled framework, perhaps incorporating probabilistic modelling or data-driven selection criteria, to guide variable inclusion and classification. Such a framework would not only improve the reproducibility and interpretability of emulation studies but also reduce subjective bias. In our study, we worked closely with clinical experts to define confounders and ensure alignment with domain knowledge, reflecting a pragmatic approach in the absence of universally accepted standards.

Our method also has several other limitations. First, our analysis is restricted to a single clinical trial, and specifically to a trial with a negative primary outcome. This narrow scope may introduce selection bias and limit the generalizability of our findings. Second, the analysis was conducted exclusively on Mayo Clinic data, which may differ in demographic or clinical characteristics from other healthcare systems, potentially affecting the external validity of our results. Third, we used the trial’s official completion date as the temporal cutoff for cohort separation. However, this may not accurately capture the true timing of the trial’s influence on clinical practice, as the dissemination of results and subsequent adoption by different institutions may occur after publication and vary across sites. Future work should consider multiple trials across a range of outcomes, incorporate diverse datasets, and explore more flexible or data-driven definitions of the transition point for clinical uptake.

## 4. Methods

Figure 3 outlines the overall workflow of our emulation study. It begins with the emulation of the WARCEF trial using real-world EHR data, where patient cohorts are constructed based on eligibility criteria. The figure highlights stratification by treatment time, before and after the trial’s completion in July 2014, to examine potential shifts in treatment effect.

**Figure 3.**
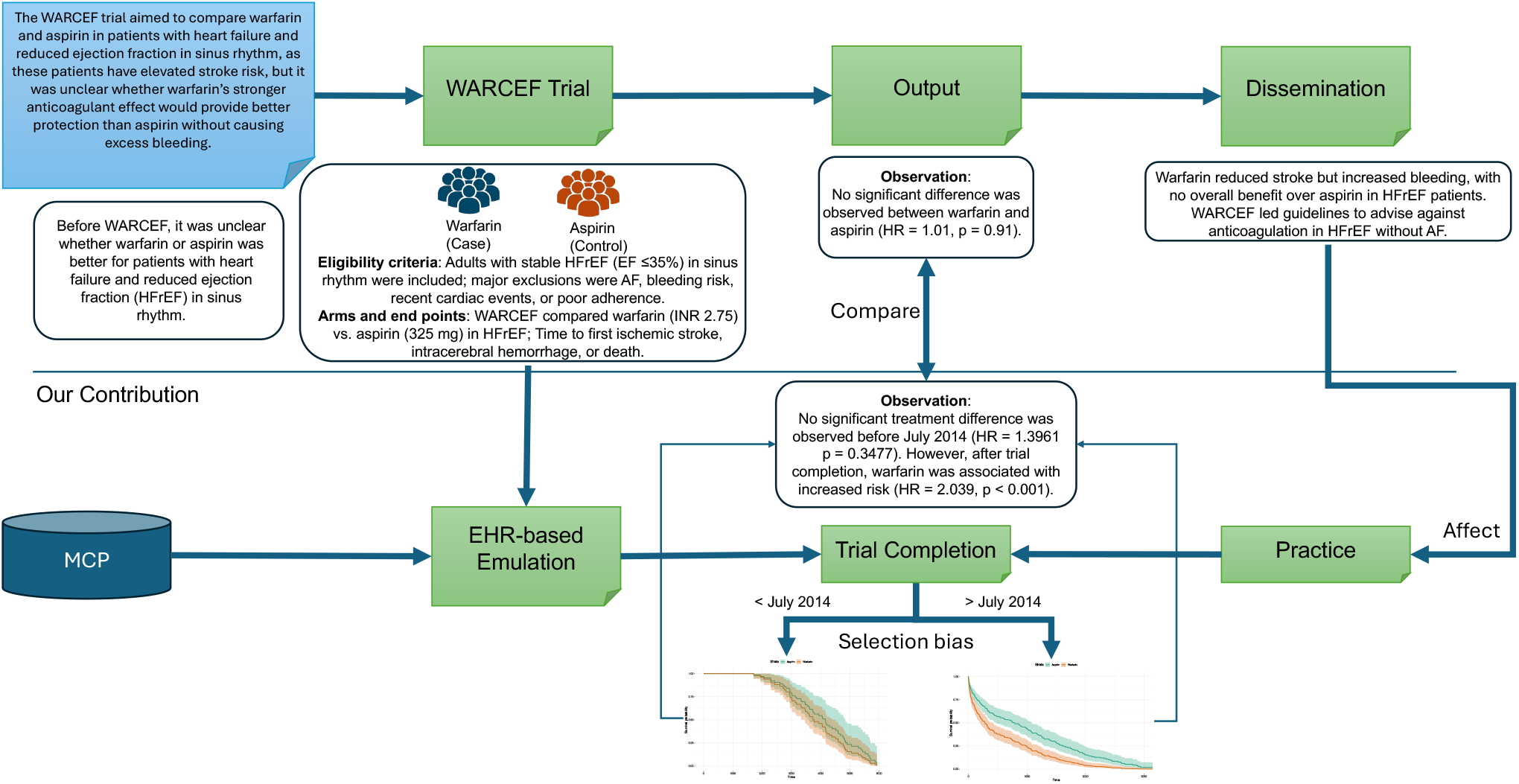
Overview of the proposed method. This figure summarizes the WARCEF trial and its EHR-based emulation. The WARCEF trial evaluated Warfarin versus Aspirin in patients with HFrEF in sinus rhythm, finding no significant difference in overall outcomes: Warfarin reduced ischemic stroke but increased bleeding, leading to guideline recommendations favouring Aspirin. To emulate the trial, real-world EHR data were used to construct a comparable patient cohort based on WARCEF’s inclusion and exclusion criteria. The emulation revealed a key observation: before July 2014 (trial completion), the difference between Warfarin and Aspirin was not statistically significant, aligning with the trial’s findings; however, after July 2014, a significant difference emerged, suggesting that the dissemination of WARCEF influenced clinical practice and outcomes in the broader patient population.

In our emulation, patient cohorts were constructed using the Mayo Clinic Platform based on key eligibility criteria and refined to ensure comparability between treatment groups. Patients were categorized into Warfarin and Aspirin groups. All outcomes were extracted from electronic health records using definitions aligned with prior clinical trials. In our case, we use death as the outcome. To enhance feasibility and maintain the integrity of our analysis, we employed both an ITT [19] approach and a PP [20] analysis. The ITT approach includes all participants in their originally assigned treatment groups, regardless of adherence, thereby preserving the benefits of randomization and minimizing selection bias. This method offers a realistic estimate of treatment effectiveness in typical real-world conditions. In parallel, the PP analysis was conducted to assess treatment efficacy under ideal conditions by including only participants who adhered strictly to the study protocol. While PP can provide insights into the potential maximum effect of the intervention, it may introduce bias by excluding non-adherent individuals and, therefore, may limit generalizability. By implementing both approaches, we aimed to provide a balanced evaluation of the intervention’s effectiveness and efficacy. In our case, the ITT analysis includes all patients regardless of the duration of medication use, whereas the PP analysis includes only those who took the medication for at least six years or died before their final medication date, following the WARCEF protocol standards. Patients were further stratified by treatment date, before or after July 2014, to assess the potential impact of trial completion.

Table 3 provides a detailed comparison between the traditional RCT design and our target trial emulation approach. It outlines key aspects, including study aim, eligibility criteria, treatment strategies, follow-up period, and outcome assessment. This comparison highlights the methodological differences, particularly in how the target trial emulation leverages RWD to approximate an RCT while addressing feasibility and generalizability in clinical research. To align with the original WARCEF protocol, we systematically mapped the trial’s inclusion and exclusion criteria to structured EHR variables available within the Mayo Clinic Platform. Each criterion was classified as a must-include, must-exclude, or confounder based on its interpretability, availability, and influence on treatment assignment and outcome. Among the inclusion criteria, left ventricular ejection fraction (LVEF ≤ 35%) or wall motion index (WMI ≤ 1.2) measured within three months in a stable clinical state was treated as a strict inclusion criterion and applied without exception. In contrast, other criteria such as LVEF timing beyond 3 months post-intervention (e.g., CABG, PTCA), outpatient protocol adherence, and discharge status after IV CHF were not considered due to lack of structured availability or feasibility in routine data. The Modified Rankin score (<4) was used as a confounder, acknowledging its potential association with both treatment decisions and outcomes. Similarly, recent stroke or transient ischemic attack (TIA) within 12 months and ACE inhibitor use (or substitute therapies) were treated as confounders to preserve patient volume and statistical power while adjusting for their prognostic relevance.

**Table 3.** Comparison between Randomized Clinical Trial and Our Target Trial Design.

| **Trials** | **Randomized clinical trial**<br>**(NCT00041938)** | **Our target trial design** |
| --- | --- | --- |
| Aim | Warfarin Versus Aspirin in Reduced Cardiac Ejection Fraction (WARCEF) Trial (WARCEF) | Determine whether Warfarin or Aspirin is better for preventing death or stroke in patients with poor heart function |
| Eligibility criteria | **Inclusion criteria:**<br>Cardiac EF $\leq 35\%$ by radionuclide ventriculography, left ventriculography or quantitative echocardiographic measurement or an echocardiographic Wall Motion Index of $\leq 1.2$, within three months of enrollment. The qualifying left ventricular function measurement must be obtained at least three months after an MI, coronary bypass grafting, PTCA, and at least one month after pacemaker insertion. Patients scheduled for mitral valve repair should have qualifying echo after surgery.<br>Modified Rankin score $\leq 4$.<br>Patient must be taking ACE inhibitors. If intolerant of ACE inhibitor, patient must be on angiotensin II receptor blockers or hydralazine and nitrates.<br>Recent stroke/TIA within 12 months<br>**Exclusion criteria:**<br>The presence of any of the following embolism: chronic or paroxysmal AF, mechanical valve, endocarditis, intracardiac mobile or pedunculated thrombus, and valvular vegetation.<br>Cardiac surgery/angioplasty/MI within 3 months.<br>Cerebral/systemic hemorrhage in past year<br>Age $< 18$ | **Criteria strictly included:**<br>Patients with Cardiac EF $\leq 35\%$ ($\leq 35\%$ is used as a covariate in analysis) by radionuclide ventriculography, left ventriculography or quantitative echocardiographic measurement or an echocardiographic Wall Motion Index of $\leq 1.2$ ($\leq 1.2$ is used as a covariate in analysis), within three months of enrollment. The qualifying left ventricular function measurement must be obtained at least three months after an MI, coronary bypass grafting, PTCA, and at least one month after pacemaker insertion.<br>**Criteria used as confounders:**<br>Modified Rankin score $\leq 4$ or Rankin score $\leq 4$<br>Patients must be taking ACE inhibitors. If intolerant of ACE inhibitor, patient must be on angiotensin II receptor blockers or hydralazine and nitrates.<br>Recent stroke/TIA within 12 months<br>**Criteria strictly excluded:**<br>The presence of any of the following embolism: chronic or paroxysmal AF, mechanical valve, endocarditis, intracardiac mobile or pedunculated thrombus, and valvular vegetation.<br>Cardiac surgery/angioplasty/MI within 3 months.<br>Cerebral/systemic hemorrhage in past year.<br>Age $< 18$<br>**Criteria used as confounders:** |

Table 3. (Continued)
| Trials | Randomized clinical trial<br>(NCT00041938) | Our target trial design |
| --- | --- | --- |
| Eligibility criteria | Cyanotic congenital heart disease, Eisenmenger's syndrome.<br>Decompensated heart failure.<br>Contraindications (e.g., GI bleed, INR>1.3, low platelets, Hct<30).<br>NSAID use.<br>Alcohol/substance abuse, gait instability.<br>Severe liver impairment.<br>Severe uncontrolled hypertension.<br>Positive stool guaiac (non-hemorrhoidal).<br>Creatinine >3.0.<br>Need for heparin/antiplatelet agent.<br>Dementia or psychiatric or physical problem that prevents the patient from following an outpatient program reliably.<br>Comorbid conditions that may limit survival to less than five years.<br>Pregnancy, or female of childbearing potential who is not sterilized or is not using a medically accepted form of contraception* (see procedure manual). *A pregnancy test is required for all women of childbearing age.<br>Hospitalization for new diagnosis of onset CHF within the past one month or carotid endarterectomy or pacemaker insertion within the past one month prior to randomization. | Cyanotic congenital heart disease, Eisenmenger's syndrome.<br>Decompensated heart failure.<br>Contraindications (e.g., GI bleed, INR>1.3, low platelets, Hct<30).<br>NSAID use.<br>Alcohol/substance abuse, gait instability.<br>Severe liver impairment.<br>Severe uncontrolled hypertension.<br>Positive stool guaiac (non-hemorrhoidal).<br>Creatinine >3.0.<br>Need for heparin/antiplatelet agent<br>Dementia or psychiatric or physical problem that prevents the patient from following an outpatient program reliably.<br>Comorbid conditions that may limit survival to less than five years.<br>Pregnancy, or female of childbearing potential who is not sterilized or is not using a medically accepted form of contraception* (see procedure manual). *A pregnancy test is required for all women of childbearing age.<br>Hospitalization for new diagnosis of onset CHF within the past one month or carotid endarterectomy or pacemaker insertion within the past one month prior to randomization. |
| Treatment strategies | Treatment 1: Aspirin (325 mg per day).<br>Treatment 2: Warfarin [International Normalized Ration (INR) 2.5-3.0; target INR = 2.75]. | Cohorts selected with medication administered:<br>Aspirin (Dose is used as a covariate in analysis).<br>Warfarin (INR is used as a covariate in analysis). |
| Treatment assignment | Randomized | Propensity score matching and inverse probability of treatment weighting. |
| Follow-up period | 2002.10 -- 2011.08 | Time Zero: First occurrence of all eligibility criteria satisfied.<br>Follow-up: Up to 6 years from Time Zero |
| Outcome | Composite Endpoint of Ischemic Stroke, Intracerebral Hemorrhage, or Death. | Endpoint of Death. |

Regarding exclusion criteria, several conditions were handled as strict exclusion filters, including: (1) Chronic/paroxysmal atrial fibrillation, mechanical valve, endocarditis, thrombus, or vegetation; (2) Cardiac surgery, angioplasty, or myocardial infarction within 3 months; (3) Cerebral or systemic hemorrhage within the past year; (4) Age < 18 years. These were directly excluded to ensure fidelity to trial conditions and minimize major sources of heterogeneity. Other clinical conditions that may influence both treatment and prognosis were modelled as confounders rather than hard exclusions. These included: (1) Cyanotic congenital heart disease and Eisenmenger’s syndrome; (2) Decompensated heart failure; (3) Contraindications to treatment (e.g., GI bleeding, abnormal INR or platelet counts, low hematocrit); (4) Use of NSAIDs; (5) Severe liver impairment, uncontrolled hypertension, elevated creatinine levels, and positive stool guaiac tests; (6) Pregnancy, dementia, and psychiatric/physical disability; (7) History of substance abuse, short life expectancy, lack of contraception, or recent CHF-related procedures. While the protocol called for strict exclusion of these patients, they were instead retained and statistically adjusted for to avoid excessive sample loss and maintain analytic robustness. Finally, certain exclusion criteria, such as “need for heparin/antiplatelet agent”, were retained in the cohort but treated as confounders due to the extremely small number of affected patients, making exclusion infeasible. A small number of criteria, such as “life expectancy <5 years” or contraception status, were not considered due to their absence or ambiguity in structured EHR data.

## Data Availability

All data produced in the present study are not available publically

## Acknowledgments

This work is supported by a grant from the National Institute of Health (NIH) NIGMS (R00GM135488), NIH R01AG084236, and Mayo Clinic Department Chair funding.

## Contributions

NZ and CT conceptualized and supervised the whole study. NZ led and managed this project. XL, SR, and XH contributed to data processing, emulation design, and experiment. JF, PF, SB, and RL facilitated access to the MCP data and planform. KL provided the domain expertise for the conduction of the emulation. NZ and XL composed the initial draft of the report and all authors reviewed the manuscript. The final manuscript was approved by all authors.

## Data and Code Availability

This study involves analysis of de-identified Electronic Health Record (EHR) data via Mayo Clinic Platform Discover. In accordance with the Code of Federal Regulations, 45 CFR 46.102, the noted activity does not require IRB review. Data shown and reported in this manuscript has been extracted from the EHR using an established protocol for data extraction, aimed at preserving patient privacy. The data has been determined to be de-identified pursuant to an expert’s determination in accordance with the HIPAA Privacy Rule. Any data beyond what is reported in the manuscript, including but not limited to the raw EHR data, cannot be shared or released due to the parameters of the expert determination to maintain the data de-identification. Contact corresponding authors for additional details regarding Mayo Clinic Platform Discover.

## Appendix

**supplementary Figure 1.**
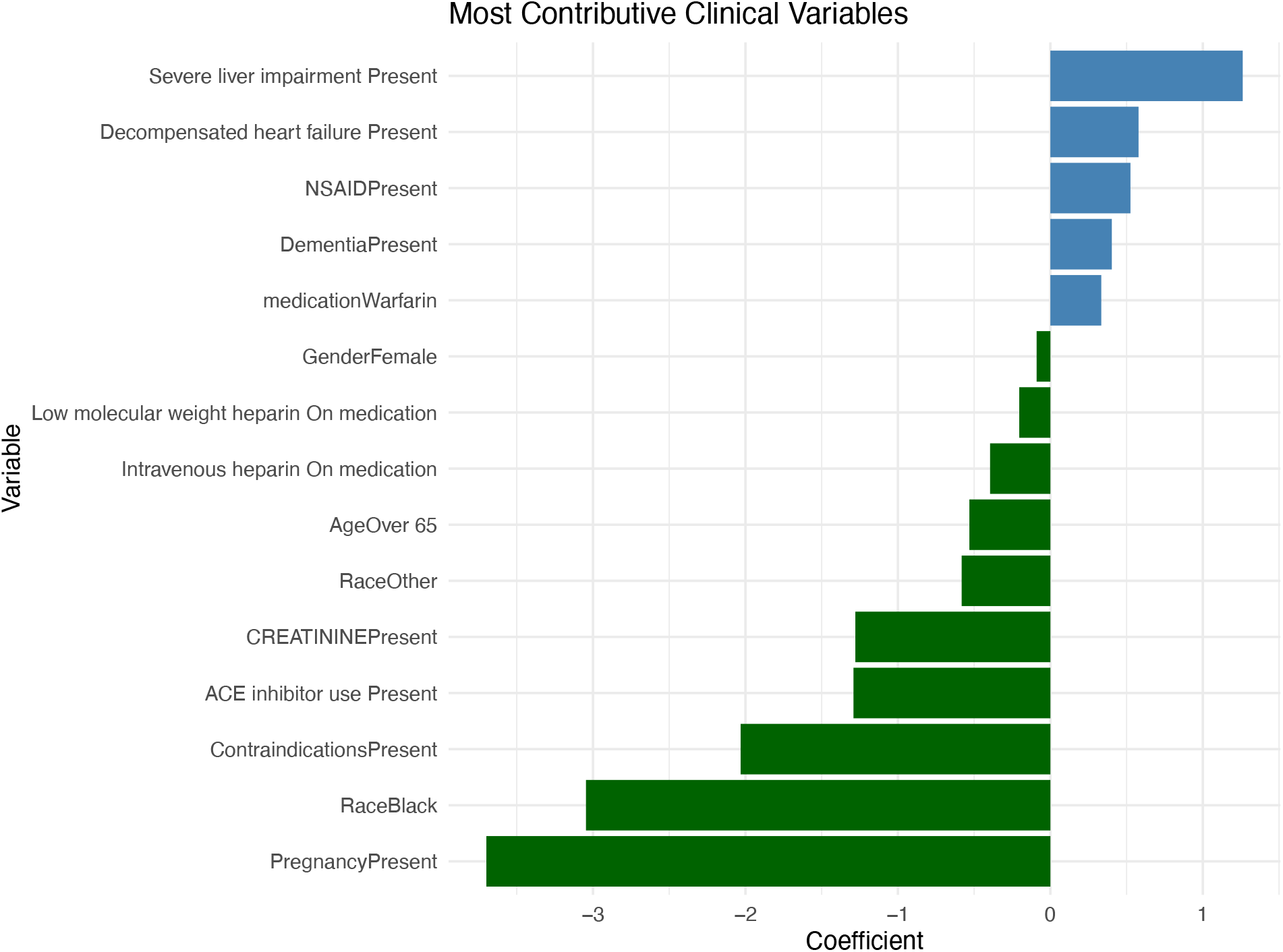
Confounders Coefficient of ITT before 2014. This figure displays the most influential clinical variables contributing to the outcome in the ITT analysis conducted prior to 2014. The horizontal bar plot ranks variables by the magnitude of their regression coefficients, with variable names listed on the y-axis and their corresponding coefficients on the x-axis. Positive coefficients indicate a stronger association with increased risk or likelihood of the outcome, whereas negative coefficients reflect a protective effect or decreased risk. Among the top positive contributors are variables such as “Severe liver impairment Present” and “Decompensated heart failure Present”, suggesting these factors are associated with elevated outcome risk. Similarly, “NSAIDPresent” appear high on the list, reinforcing their clinical importance in the prediction model. On the other hand, several variables demonstrate negative coefficients, indicating a potential protective association. Notably, “PregnancyPresent”, “RaceBlack”, and “ContraindicationsPresent” have the most negative coefficients, suggesting these factors are linked to reduced risk or lower predicted outcomes in this specific cohort and timeframe. Other negatively associated features include “ACE inhibitor use Present” and “CREATININEPresent”, which may reflect their influence in moderating outcome severity. The transition from negative to positive coefficients from top to bottom in the plot highlights the shift in influence among clinical variables. This visualization provides important insight into which factors most strongly affect patient outcomes before 2014, with variables related to liver function, age, and comorbid conditions playing a prominent role in risk stratification.

**supplementary Figure 2.**
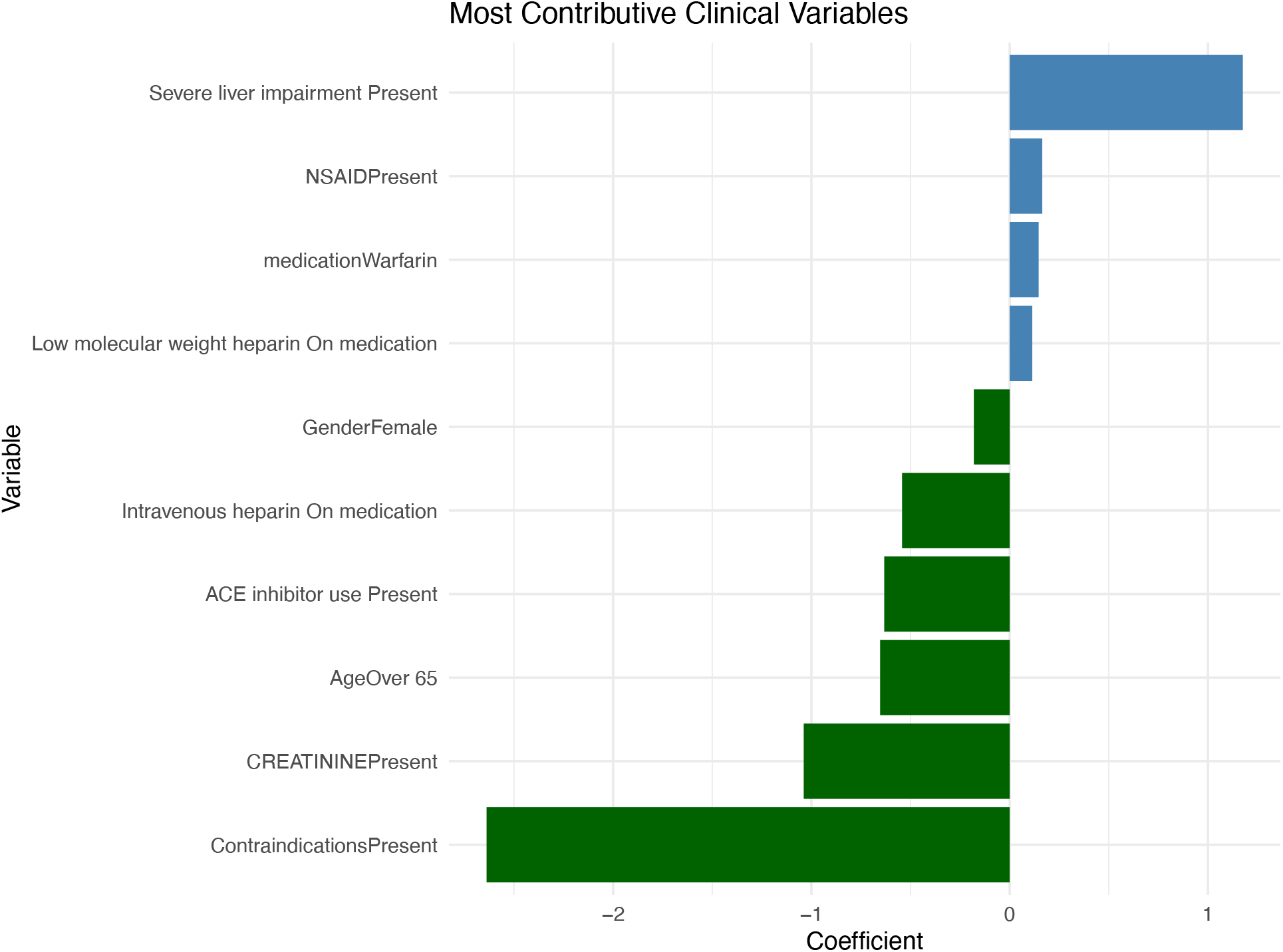
Confounders Coefficient of PP before 2014. This figure presents the most contributive clinical variables associated with outcomes in the PP analysis prior to 2014. The horizontal bar chart displays clinical variables on the y-axis and their corresponding coefficient values on the x-axis. Positive coefficients indicate an increased risk or likelihood of the outcome, while negative coefficients suggest a protective effect or reduced risk. Among the top positive contributor is “Severe liver impairment Present”, is known to be clinically relevant indicators of higher patient risk. Other notable variables with positive associations include “NSAIDPresent”, “medicationWarfarin”, and “Low molecular weight heparin On medication”, indicating that medication use and demographic factors may also influence outcomes in this population. Conversely, several variables demonstrate negative coefficients, pointing to a potential protective effect. Most prominently, “ContraindicationsPresent” has the most negative coefficient, followed by smaller negative contributions from “ACE inhibitor use Present”, “Intravenous heparin On medication”, and “GenderFemale”. These findings may reflect treatment selection patterns or confounding by indication, where certain interventions are linked with lower observed outcome risk under the per-protocol setting. This visualization offers insights into the clinical factors most associated with outcome variation before 2014, with organ dysfunction, age, and contraindications emerging as influential variables within the PP analytic framework.

**supplementary Figure 3.**
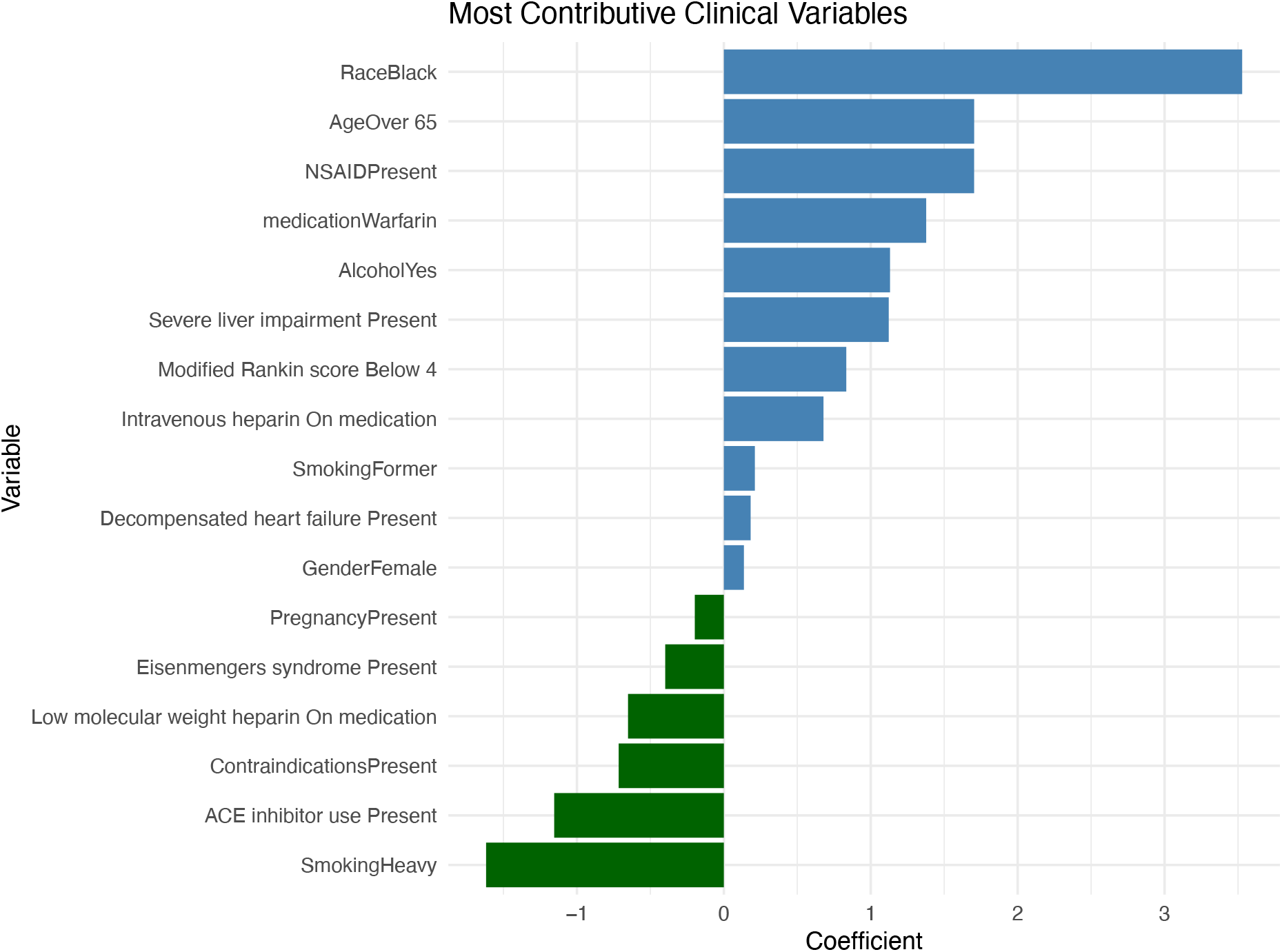
Confounders Coefficient of PP after 2014. This figure presents the most contributive clinical variables affecting the outcome in the PP analysis conducted after 2014. The horizontal bar plot ranks variables by the magnitude of their coefficients, with variable names listed on the y-axis and corresponding coefficient values on the x-axis. Positive coefficients indicate a stronger association with increased risk or higher probability of the outcome, while negative coefficients reflect a potential protective effect or reduced risk. The most significant positive contributors include “RaceBlack”, “AgeOver 65”, and “NSAIDPresent”, all of which exhibit high positive coefficients, suggesting a strong influence on adverse outcomes. Other notable features with positive associations are “medicationWarfarin” and “AlcoholYes”, reinforcing the impact of lifestyle and medication-related variables in this post-2014 cohort. Mid-level contributors include “Severe liver impairment Present”, and “Intravenous heparin On medication”, which continue to show positive associations with the outcome, albeit to a lesser extent. At the lower end of the coefficient scale, variables such as “Eisenmengers syndromePresent” and “PregnancyPresent” exhibit negative coefficients, indicating a potential protective or less harmful influence on outcomes. “GenderFemale” also appears at the bottom of the list with the smallest (most positive) coefficient. This visualization offers insight into the evolving impact of clinical variables on outcome prediction after 2014, highlighting shifts in demographic and medication-related risk factors, with particular emphasis on age, race, and treatment characteristics.

